# A rapid pharmacogenomic assay to detect *NAT2* polymorphisms and guide isoniazid dosing for tuberculosis treatment

**DOI:** 10.1101/2021.01.17.21249995

**Authors:** Renu Verma, Sunita Patil, Nan Zhang, Flora M.F. Moreira, Marize T. Vitorio, Andrea da S. Santos, Ellen Wallace, Devasena Gnanashanmugam, David Persing, Rada Savic, Julio Croda, Jason R. Andrews

**Affiliations:** Division of Infectious Diseases and Geographic Medicine, Stanford University School of Medicine, Stanford, CA, USA; Department of Bioengineering and Therapeutic Sciences, University of California, San Francisco, CA, USA; Federal University of Grande Dourados, Dourados, Brazil; Cepheid Inc., Sunnyvale, California, USA; Postgraduate Program in Infectious and Parasitic Diseases, Federal University of Mato Grosso do Sul, Mato Grosso do Sul, Brazil; Oswaldo Cruz Foundation Mato Grosso do Sul, Mato Grosso do Sul, Brazil

**Keywords:** tuberculosis, isoniazid, pharmacogenomic, molecular diagnostic, *NAT2*

## Abstract

**Rationale:** Standardized weight-based dose of anti-tubercular drugs contributes to a substantial incidence of toxicities, inadequate treatment response, and relapse, in part due to variable drug levels achieved. Single nucleotide polymorphisms (SNPs) in the N-acetyltransferase-2 *(NAT2)* gene explain the majority of interindividual pharmacokinetic variability of isoniazid (INH). However, an obstacle to implementing pharmacogenomic-guided dosing is the lack of a point-of-care assay.

**Objectives:** To develop and test a *NAT2* classification algorithm, validate its performance in predicting isoniazid clearance, and develop a prototype pharmacogenomic assay.

**Methods:** We trained random forest models to predict *NAT2* acetylation genotype from unphased SNP data using a global collection of 8,561 phased genomes. We enrolled 48 pulmonary TB patients, performed sparse pharmacokinetic sampling, and tested the acetylator prediction algorithm accuracy against estimated INH clearance. We then developed a cartridge-based multiplex qPCR assay on the GeneXpert platform and assessed its analytical sensitivity on whole blood samples from healthy individuals.

**Measurements and Main Results:** With a 5-SNP model trained on two-thirds of the data (n=5,738), out-of-sample acetylation genotype prediction accuracy on the remaining third (n=2,823) was 100%. Among the 48 TB patients, predicted acetylator types were: 27 (56.2%) slow, 16 (33.3%) intermediate and 5 (10.4%) rapid. INH clearance rates were lowest in predicted slow acetylators (median 19.3 L/hr), moderate in intermediate acetylators (median 41.0 L/hr) and highest in fast acetylators (median 46.7 L/hr). The cartridge-based assay accurately detected all allele patterns directly from 25ul of whole blood.

**Conclusions:** An automated pharmacogenomic assay on a platform widely used globally for tuberculosis diagnosis could enable personalized dosing of isoniazid.

**Summary:** This manuscript describes the development and validation of point-of-care multiplex pharmacogenomic assay to guide personalized dosing of isoniazid for treatment or prevention of tuberculosis.

## INTRODUCTION

Despite the availability of effective chemotherapeutic regimens for treatment and prevention of tuberculosis, a substantial proportion of patients experience toxicities, fail treatment or develop recurrent disease (1-3). Standardized, weight-based dosing of anti-tuberculosis treatment has been the conventional approach to therapy, despite mounting evidence that inter-individual variability in metabolism leads to highly variable drug levels (4,5). High drug levels are strongly associated with risk of toxicity, while low drug levels are a determinant of treatment failure, slow response, and emergence of drug resistance. Hepatotoxicity is the most common adverse effect, affecting up to 33% of patients receiving standard four-drug therapy (6) and leading to regimen changes in up to 10% of patients (7). This toxicity is associated with increased costs, morbidity, and occasional mortality, particularly among HIV co-infected individuals (8). Additionally, as many as 3% of new tuberculosis cases experience treatment failure, and between 6-10% relapse within 2 years (9,10). Pharmacokinetic variability to a single drug is associated with treatment failure and acquired drug resistance (11,12). One study found that individuals with at least one drug below the recommended AUC threshold had a 14-fold increased risk of poor outcomes (13)

More than 50 years after its introduction, isoniazid (INH) remains one of the major first line drugs used to treat active and latent tuberculosis infections (14,15). There has been an increasing number of genetic markers identified that predict metabolism and toxicities from various antimicrobials. INH is among the most well characterized of these, with more than 80% of its pharmacokinetic variability explained by mutations in the gene encoding arylamine N-acetyltransferase 2 (NAT2), responsible for its metabolism in the liver (16-18). The primary step in the metabolism of INH is acetylation, catalyzed by the NAT2 enzyme, resulting in the formation of acetyl-INH. The NAT2 enzyme displays genetic polymorphism, and its activity is expressed at highly variable levels. A high correlation between INH acetylator phenotype and seven most frequent SNPs in *NAT2* gene has been demonstrated by several studies (19-22). Individuals can be classified into three phenotypes—rapid, intermediate, and slow acetylators— according to whether they carry polymorphisms on neither, one, or both copies of this gene, respectively. Rapid acetylators typically have the lowest plasma INH concentrations, while slow acetylators have high concentrations (23). A worldwide population survey on *NAT2* acetylation phenotype reported that more than half of the global population are slow or rapid acetylators (18). A meta-analysis of 14 studies, comprising 474 cases and 1446 controls based on *NAT2* polymorphisms found that rapid acetylators are twice as likely to have microbiological failure and acquired drug resistance. Additionally, a significant association has been consistently observed between *NAT2* slow acetylators and the risk of anti-tuberculosis drug-induced liver injury (17). Additional meta-analyses have identified a three-to four-fold increased risk of hepatotoxicity among slow acetylators (24). A randomized trial of pharmacogenomic guided dosing for tuberculosis treatment found that, compared with standard dosing, it reduced hepatotoxicity among slow acetylators and increased treatment response at 8 weeks among rapid acetylators (25)

Despite this evidence, pharmacogenomic testing and guided treatment has not entered the mainstream of clinical practice for tuberculosis. Few clinical laboratories perform *NAT2* genotyping, which requires detection of multiple polymorphisms and testing for heterozygous allele patterns. Such testing is not widely available in resource-constrained environments where the majority of tuberculosis burden falls. To address this gap, we developed an algorithm from unphased SNP patterns, derived from globally representative genomic data, to predict INH metabolism phenotype using fewer SNPs while retaining high accuracy. Based on the SNP combination derived from this model, we further developed a prototype *NAT2* pharmacogenomic (NAT2-PGx) assay on a commercial, automated PCR platform (GeneXpert) to detect *NAT2* polymorphisms. We demonstrate that this tool can accurately predict INH clearance rates directly from clinical samples and can be easily performed with minimal training and hands-on time.

## METHODS

### Datasets

The datasets used to develop the NAT2 classifier was obtained from the IGSR (International Genome Sample Resource, 1000 genomes project) and a meta-analysis by Sabbagh et. al (18, 26). Population information on the combined dataset is provided in **Supplementary Table 1**.

### *NAT2* acetylator phenotype prediction classifier

Phased genomes from 8,561 individuals were used and haplotypes were labeled based on seven most frequent SNPs (both synonymous and non-synonymous) which are reported to affect the acetylation on INH - rs1801279 (191G>A), rs1041983 (282C>T), rs1801280 (341T>C), rs1799929 (481C>T), rs1799930 (590G>A), rs1208 (803A>G) and rs1799931 (857G>A) (19) in the *NAT2* gene, following an international consensus nomenclature (27). We then constructed an unphased dataset containing only information on whether each sample was wild type for both alleles, homozygous variant for both alleles, or heterozygous. We stratified the dataset by geographic region and then drew a random sample from two-thirds of each stratum for a training set and one third for an out-of-sample test set, to ensure geographic representativeness in the training and test sets. We trained random forest models on the training set using the *caret* package in R and assessed classification performance on the held-out test set. We began with a 7 SNP model and eliminated SNPs in sequential models according to the lowest variable importance factor.

### Ethics statement

The clinical study was approved by the institutional review boards of the Stanford University School of Medicine and Federal University of Grande Dourados. All participants were over the age of 18 and provided written informed consent. For assay optimization and validation on GeneXpert cartridge, anonymized blood samples from healthy individuals were obtained from Stanford blood center.

### Sample collection

Sputum and plasma samples from 48 newly diagnosed patients with active pulmonary tuberculosis were collected at the Federal University of Grande Dourados, Brazil. All participants were treated with standardized, weight-based doses of isoniazid, rifampicin, pyrazinamide and ethambutol. Plasma samples were collected at 1 hour and 8 hours after the first dose and after 1 hour on day 14. Plasma drug concentrations for isoniazid and acetyl-isoniazid were quantified by high-performance liquid chromatography coupled to tandem mass spectrometry (HPLC-MS) as previously described (28).

### Pharmacokinetic analysis of INH clearance in tuberculosis patients

The population PK analysis was performed using the non-linear mixed effects modeling approach using NONMEM (version 7.4.3; ICON plc, Gaithersburg, MD, USA), PsN and R-based Xpose (version 4.7 and higher) (29,30). One-compartment model with a first-order absorption with a lognormal distribution for inter-individual variability (IIV) on different PK parameter(s) as well as an additive and/or proportional model for the residual error were tested for the base model selection. Mixture models with two or three subpopulations representing different clearance rate were then evaluated. The first-order conditional estimation with interaction method (FOCEI) was applied and the model-building procedure and model selection was based on the log-likelihood criterion (the difference in the minimum OFV between hierarchical models was assumed to be Chi-square distributed with degrees of freedom equal to the difference in the number of parameters between models), goodness-of-fit plots (e.g. relevant residuals against time randomly distributed around zero), and scientific plausibility of the model. Visual predictive check was conducted to evaluate whether the final model with estimated fixed-effect parameters and covariates adequately describe data.

### Sputum processing and host DNA extraction

Spontaneously expectorated sputum samples from confirmed pulmonary tuberculosis patients were collected in 10mL of guanidine thiocyanate (GTC) solution. The samples were needle sheared and centrifuged at 3000 rpm for 30 min. The supernatant was collected in Trizol LS and host DNA was extracted from the supernatant using a manual extraction method described previously (31).

### Primers and probes for melt curve analysis

We first developed single-plex melt curve assays based on molecular beacon probe chemistry for five NAT2 polymorphisms. Using same primer and probe sequences, we further developed multiplex NAT2-PGx assay compatible on GeneXpert platform. Three sets of primers and five molecular beacon probes spanning the *NAT2* gene were used. Primers and probes were designed using Beacon Designer (Premier Biosoft International, Palo, CA; version 8). A list of primers and probes sequences with their corresponding fluorophores and quenchers is provided in **Supplementary Table 2**.

### NAT2-genotyping on pulmonary TB samples

Host genomic DNA extracted from 48 sputum samples from TB positive patients was used to perform single-plex qPCR assays developed in-house. The assays were validated using TaqMan commercial genotyping assays (NAT2 TaqMan® SNP Genotyping Assays, Applied Biosystems). The genotyping was performed on StepOne Plus Real Time PCR (Applied Biosystems). The NAT2-genotypes derived from the assays were used to predict INH acetylator phenotype by our 5-SNP model.

### Automated NAT2-PGx Multiplex PCR and melt curve analysis

We combined the five single-plex NAT2 melt curve assays validated on TB samples into one and developed a multiplex assay (NAT2-PGx) on the FleXible Cartridge (Flex cart-01, Cepheid) platform. Flex cart allows automated DNA extraction from whole blood followed by PCR amplification and melt curve analysis to detect SNPs in *NAT2* gene in a single run. NAT2-PGx assay was performed on a GeneXpert IV instrument using GeneXpert Dx 4.8 software (Cepheid, Sunnyvale). Briefly, 100ul of whole blood was mixed with 900ul of GeneXpert lysis buffer for whole blood (Cepheid), incubated for 2 min at room temperature and loaded into sample preparation chamber of the flex cart for automated DNA extraction. A 70ul of PCR mastermix was simultaneously loaded in the PCR reaction chamber of the flex cart-01. PCR and melt conditions were optimized using mastermix prepared in house.

We validated the multiplex assay on blood samples from 20 healthy individuals. The accuracy of multiplex NAT2-PGx assay in SNP calling was subsequently validated by Sanger sequencing (see methods). We assessed analytical sensitivity of the assay and robustness to input blood volume by performing it on varying volumes of whole blood (200 ul, 100 ul, 50 ul, 25 ul) and comparing the Tm results and standard deviation for each position across blood volumes. Additional details on the qPCR run method and melt curve analysis is provided in an online data supplement.

## RESULTS

### SNP selection and development of acetylation prediction model

Complete phased data for the seven polymorphisms that define acetylation haplotypes were available for 8,561 individuals from 59 populations. The dataset contains 3,573 (41.7%) individuals with a slow genotype, 3,428 (40.0%) individuals with an intermediate genotype, and 1,560 (18.2%) individuals with a rapid genotype (See **Table 1**). The highest proportion of rapid acetylators were in East Asia (40%), and three regions had prevalence of slow acetylator phenotypes over 50% (Central and South Asia, Europe and North Africa). We used these phased allele data to select SNPs for inclusion in an assay measuring unphased SNPs. Using a random forest model trained on two thirds of the data (n=5,738), out-of-sample phenotype prediction accuracy from unphased data on the remaining one third (n=2,823) was 100% for models using 7, 6 or 5 SNPs. With 4 SNPs, prediction accuracy was 98.0% (95% CI: 97.4-98.5%), and a 3 SNP model had similar performance (98.0%; 95% CI: 97.4-98.4%) (**Table 2**). However, both of these models performed poorly on data from Sub-Saharan Africa (4 SNP model accuracy: 82.5%, 95% CI: 78.1-86.4%); 3 SNP model accuracy: 81.3%, 95% CI: 76.8-85.3%). Based on these results, we selected the 5 SNP model (191G>A, 282C>T, 341T>C, 590G>A and 857G>A) to take forward for clinical validation and diagnostic development.

**Table 1.** Summary of populations included in genomic analysis and their acetylation genotypes.

| Region | Number of individuals | Acetylation Genotype, n (%) |  |  |
| --- | --- | --- | --- | --- |
|  |  | Slow | Intermediate | Rapid |
| Americas | 1,112 | 432 (39%) | 463 (42%) | 217 (20%) |
| Central and South Asia | 588 | 355 (60%) | 198 (34%) | 35 (6%) |
| East Asia | 2,308 | 340 (15%) | 1049 (45%) | 919 (40%) |
| Europe | 3,458 | 1966 (57%) | 1249 (36%) | 243 (7%) |
| North Africa | 44 | 30 (68%) | 10 (23%) | 4 (9%) |
| sub-Saharan Africa | 1,051 | 450 (43%) | 459 (44%) | 142 (14%) |
| Total | 8,561 | 3573 (42%) | 3428 (40%) | 1560 (18%) |

**Table 2.**
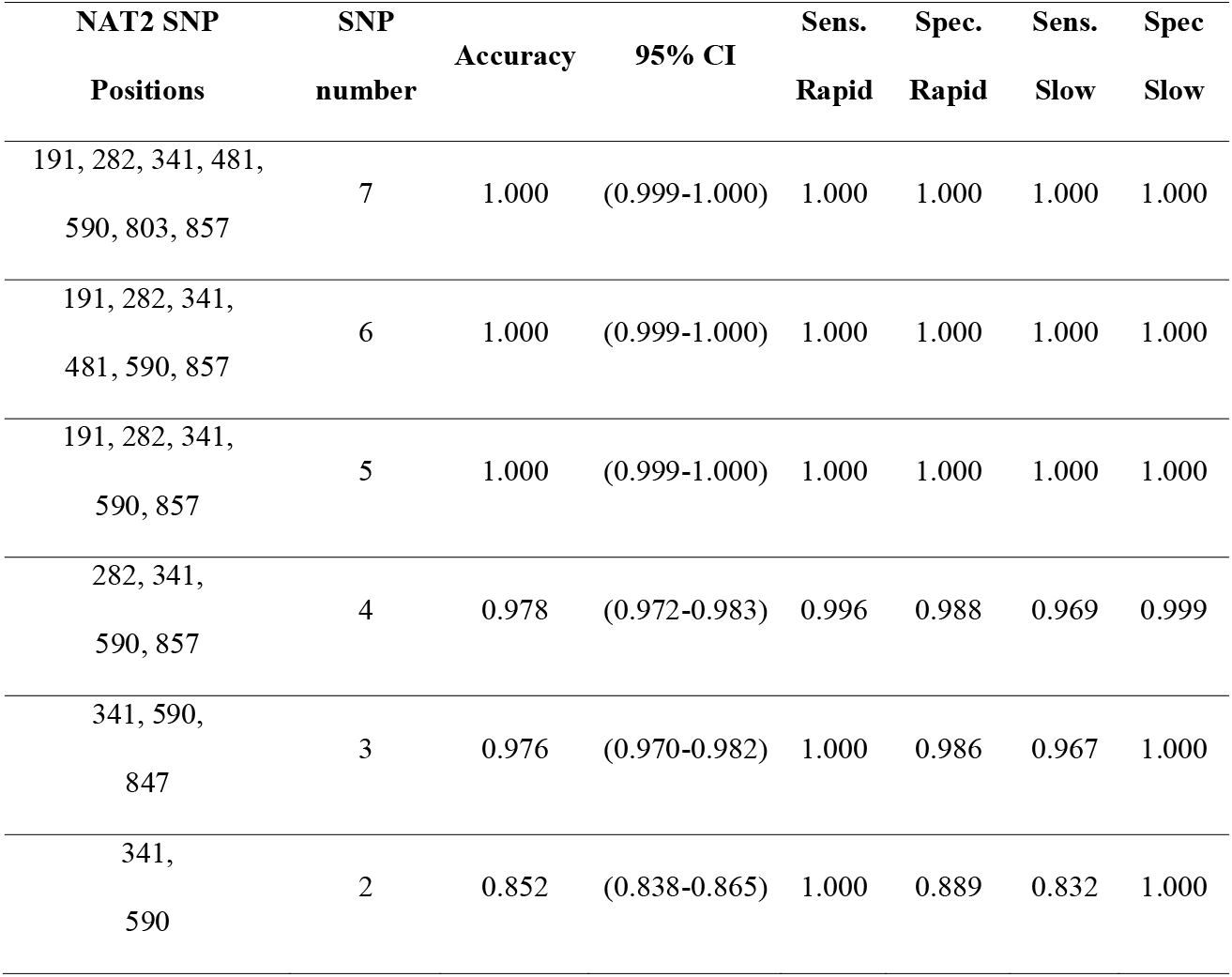
Out-of-sample prediction accuracy of unphased NAT2 SNP data for acetylation phenotype in random forest models. Models were trained with 5,738 individuals and tested on 2,823 individuals. Sens: sensitivity. Spec: specificity.

### Genotype correlation with isoniazid clearance in patients with tuberculosis

We enrolled a cohort of 48 patients with newly diagnosed pulmonary tuberculosis and collected plasma at 1 hour and 8 hours after dose on day 1 and at 1 hour after dose on day 14. To detect five *NAT2* polymorphisms identified by our classifier, we performed single-plex melt curve qPCR assays developed in-house using host DNA extracted from sputum samples. Additionally, we used commercial 7-SNP single-plex genotyping assays and compared the results with 5-SNP single-plex PCR to validate the melt curve accuracy in SNP detection. There was 100% concordance in terms of SNP detection between single-plex melt curve and commercial 7-SNP assays. Of the 48 individuals for whom *NAT2* genotypes were profiled, DNA-probe hybrid melting temperature difference (ΔTm) (°C) between wild-type and mutant alleles for positions 191, 282, 341, 590 and 857 were found to be 4.38, 4.04, 2.40, 3.63 and 3.68 respectively. Both mutant and wild type probes had a minimum 2.40°C Tm difference which allowed SNP calling with high accuracy **(Table 3)**.

**Table 3.** Melting temperature (Tm) values for five *NAT2* polymorphisms derived from DNA-probe hybrid melts using single-plex assays validated on 48 pulmonary TB patients.

|  | Mutant | Wild type | Het | Mutant | Wild type |  |
| --- | --- | --- | --- | --- | --- | --- |
| NAT2 SNP position | Total samples analyzed | Total samples analyzed | Total samples analyzed | T<sub>m</sub> ± SD | T<sub>m</sub> ± SD | ΔT<sub>m</sub> (WT-MT) |
| 191 | 0 | 44 | 5 | 66.3 ± 0.28 | 61.92 ± 0.68 | 4.38 |
| 282 | 6 | 24 | 19 | 62.81 ± 0.40 | 66.85 ± 0.34 | 4.04 |
| 341 | 12 | 23 | 14 | 66.30 ± 0.48 | 68.70 ± 0.38 | 2.40 |
| 590 | 0 | 35 | 14 | 68.0 ± 0.28 | 64.37 ± 0.28 | 3.63 |
| 857 | 1 | 41 | 7 | 66.4 ± 0.11 | 62.72 ± 0.29 | 3.68 |

SD: Standard deviation, ΔT<sub>m</sub>: DNA-probe hybrid melting temperature difference, WT: Wild type, MT: mutant

We further predicted phenotypes from 5-SNP using the algorithm described above as well as a publicly available tool (NAT2Pred) (32), which uses a 6 SNP model that excludes rs1801279 (191G>A) and includes other two sites from 7-SNP model-rs1799929 (481C>T) and rs1208 (803A>G) when compared with our 5-SNP model. Among the 48 participants, predicted acetylator types from the 5 SNP assay were: 27 (56.2%) slow, 16 (33.3%) intermediate and 5 (10.4%) rapid. NAT2Pred classified 4 samples as intermediate that were classified as rapid (n=1) or slow (n=3) by the 5 SNP classifier. Among those classified as slow by the 5 SNP classifier and intermediate by NAT2Pred, acetyl-INH to INH ratios at 8 hours were 0.61, 0.38, 0.41, consistent with slow acetylation (median: 0.76, range 0.36-1.55) rather than intermediate acetylation (median 6.67, range 3.32-22.21) and suggesting misclassification by NAT2Pred. The sample classified as intermediate by NAT2PRed and rapid by the 5 SNP classifier had an acetyl-INH to INH ratio of 9.8, which fell between the median values, and within both ranges, for intermediate and rapid acetylators (range 8.09 - ∞) (**Supplementary Table-3**). Phenotypes predicted by the 5 SNP classifier were strongly predictive of INH acetylation and clearance (**Figure 1a and 1b**). INH clearance rates were lowest in slow acetylators (median 19.3 L/hr), moderate in intermediate acetylators (median 41.0 L/hr) and highest in fast acetylators (median 46.7 L/hr).

**Figure 1.**
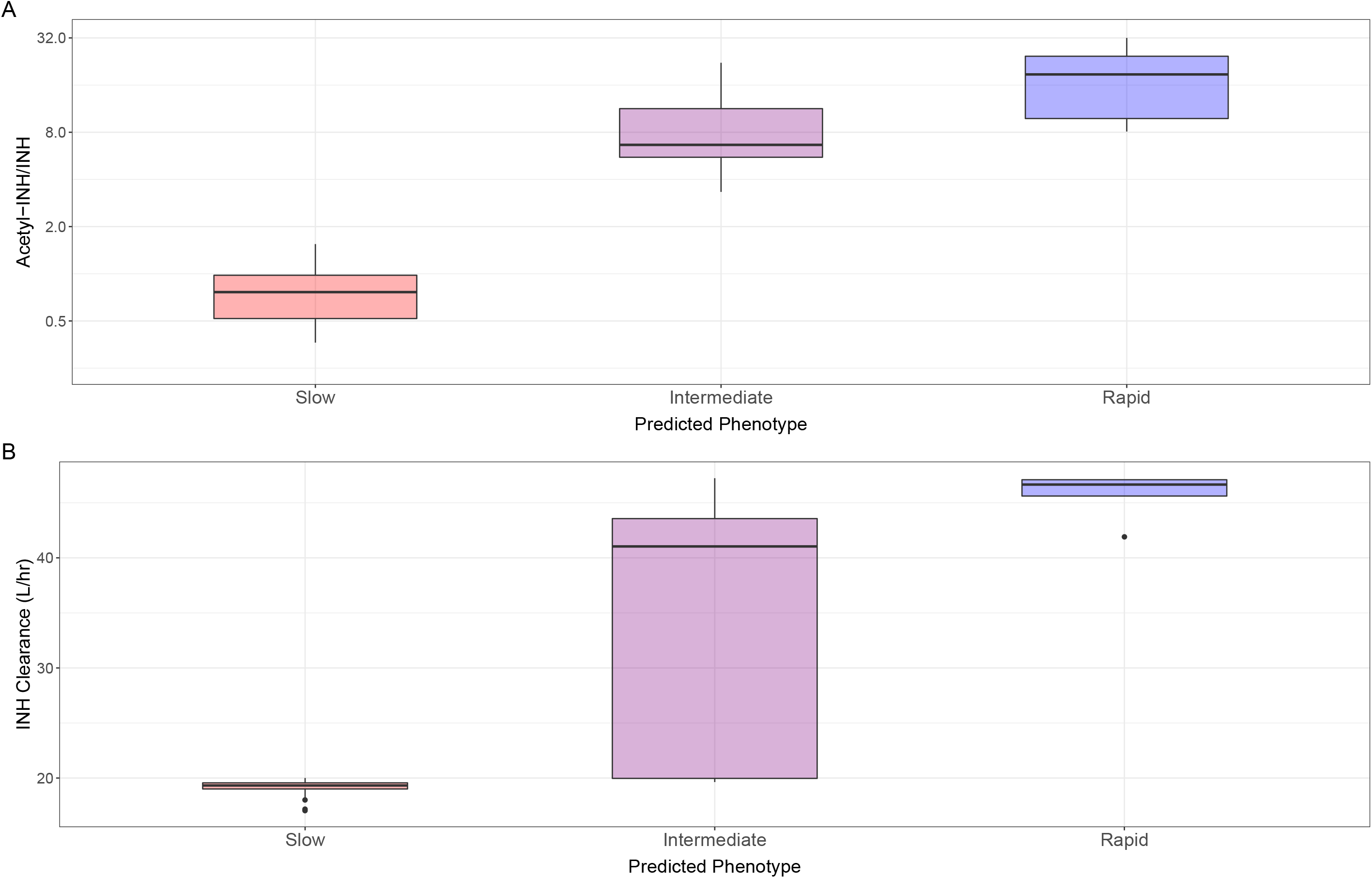
Predicted *NAT2* phenotype from sputum samples and associated acetylation ratio and isoniazid clearance rates from patients receiving tuberculosis treatment. The (a) 8 hour acetyl-INH to INH ratio and (b) isonazid clearance rates, according to acetylation phenotype predicted from 5 SNPs, measured in sputum samples from 48 patients receiving treatment for active tuberculosis.

### Development of an automated pharmacogenomic assay

We validated the NAT2-PGx assay on 20 whole blood samples from healthy individuals. The hands-on time for the cartridge preparation per sample was 5 minutes followed by 140 minutes for overall run that included automated DNA extraction (**Figure 2**). Mutant, wild-type and heterozygous alleles were manually called based on peak patterns and Tm values detected in melt curves. Negative derivative transformed melt curves from five *NAT2* gene polymorphisms are shown in **Figure 3**. The assay detected all polymorphisms with 100% accuracy (average SD in Tm across all probes = 0.34°C) compared with Sanger sequencing. The *NAT2* genotypes corresponding to 20 blood samples covered all three categories - mutant, wild-type and heterozygous for five *NAT2* positions except for NAT2-191 for which all samples were all wild-type. The 191G>A mutation is highly prevalent in African and African-American populations and is less common in other populations (33). Among the 20 samples, predicted acetylator types using the 5-SNP classifier were: 8 (40%) slow, 10 (50%) intermediate and 2 (10%) rapid (**Supplementary Table 4a**).

**Figure 2.**
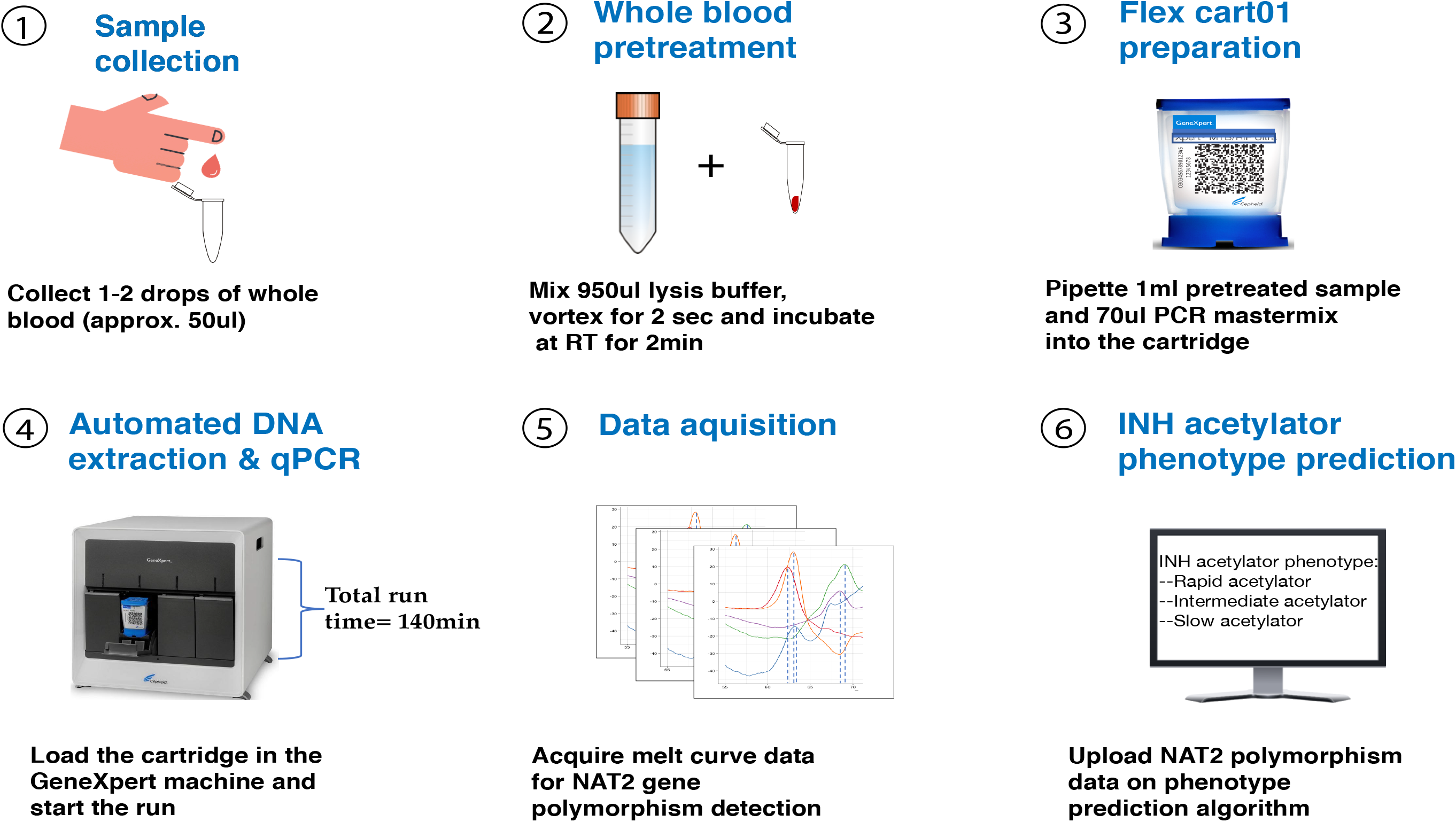
Schemata for the automated NAT2 Pharmacogenomic assay. 1-2 drops of blood is collected in an Eppendorf tube and mixed with lysis buffer to a total of 1 ml, which is then loaded onto a GeneXpert Flex01 cartridge and placed into a GeneXpert instrument for automated DNA extraction, qPCR and meltcurve analysis. Allele patterns for each of the 5 SNPs are determined by Tm analysis, and the resulting data are used to predict acetylator phenotype.

**Figure 3.**
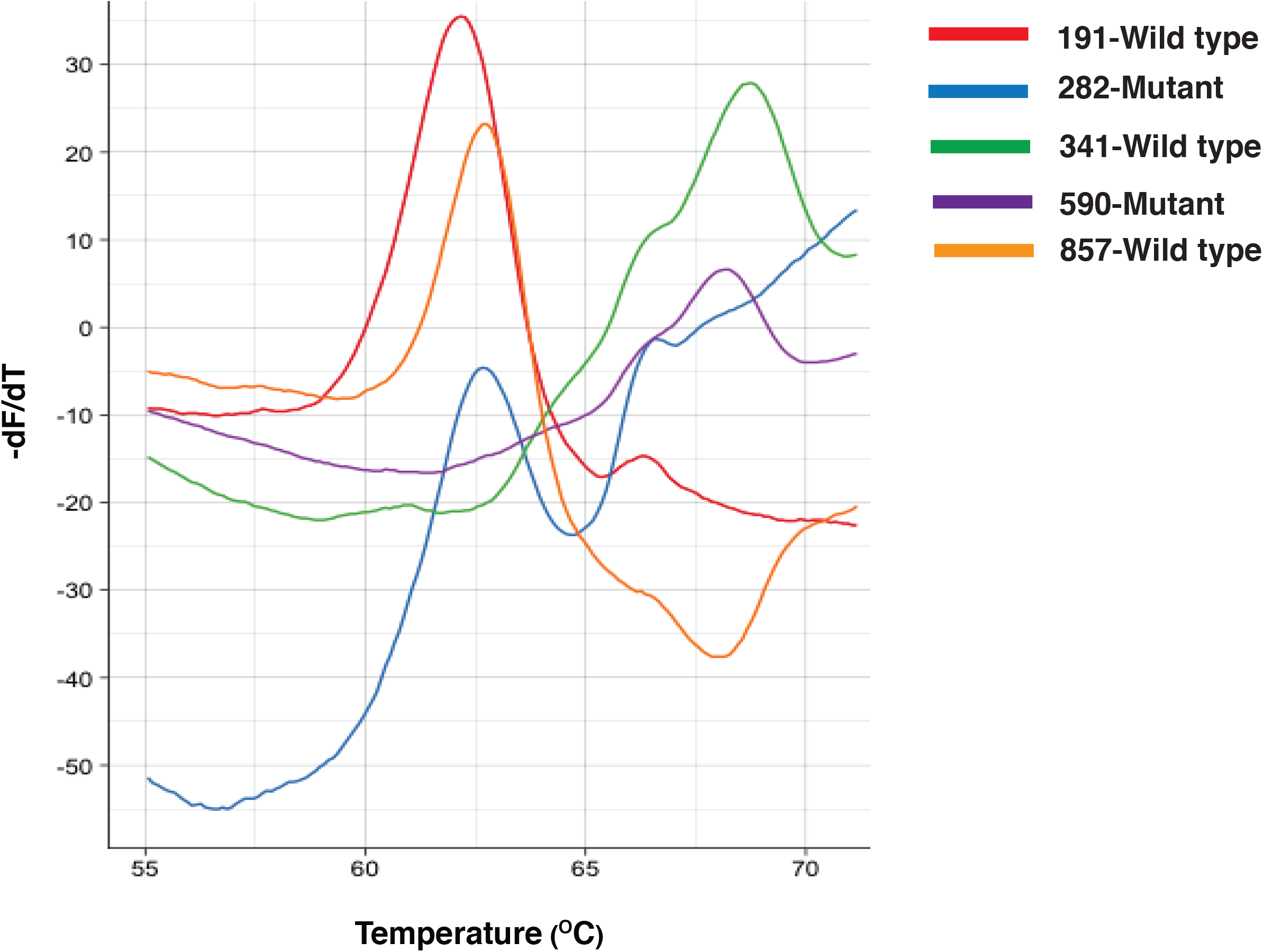
Negative derivative transformed melt curves for the five NAT2 gene polymorphisms. The shift in melt curve temperature is observed during a nucleotide exchange. Molecular beacon probes are first hybridized and then melted off of their *NAT2* target amplicon. The melt curves indicate wild type alleles at positions 191(red), 341(green) and 857 (orange); and mutant alleles at positions 282(Blue) and 590(Purple).

We further assessed the analytic performance of the NAT2-PGx assay at lower sample volumes. Our assay could accurately detect all melt peaks with as low as 25ul of sample volum**e**. The variability in Tm from five *NAT2* probes for sample volumes 200ul-25ul is shown in **Figure 4**. *NAT2* polymorphisms were accurately detected at all volumes (**Supplementary Table 4b)** and demonstrated robustness to variation in input volume.

**Figure 4.**
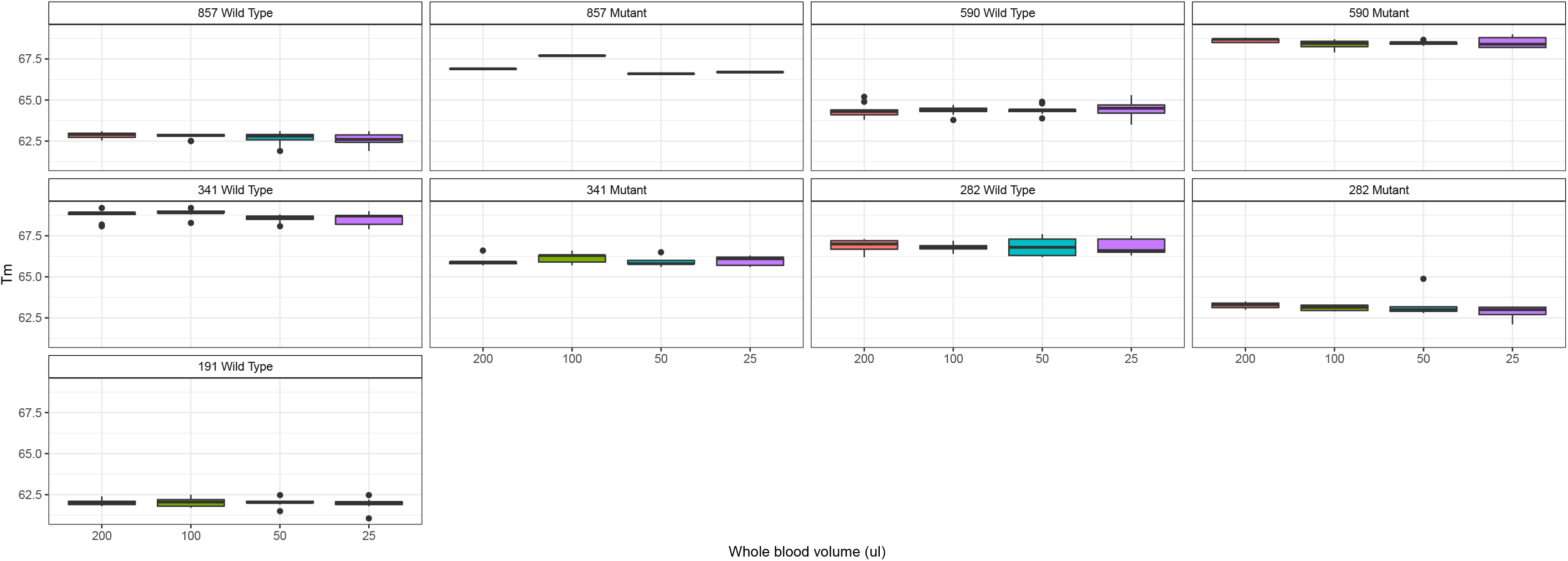
Effect of whole blood sample volume on melting temperature for wild type and mutant alleles at 5 positions in *NAT2*: *NAT2* polymorphisms were accurately detected at all volumes with sufficient different in melting temperature (Tm) to distinguish wild type from mutant alleles. None of the individuals in this dataset had mutations at position 191.

## DISCUSSION

Despite availability of effective treatment for drug-sensitive tuberculosis, a substantial proportion of population encounters drug associated toxicity or treatment failure, much of which could be averted through dosing guided by genetic markers of drug metabolism (34). We previously found that pharmacogenomic guided dosing of isoniazid could be highly cost-effective in low- and middle-income countries (35). A major barrier to its implementation has been the lack of a simple, scalable assay that could be used at points of care where tuberculosis is treated in resource-constrained settings. To address this gap, we used globally representative genomic data to identify patterns of 5 SNPs that enable accurate prediction of isoniazid acetylator phenotype, validating this with pharmacokinetic data of patients receiving tuberculosis treatment. We then developed a prototype automated pharmacogenomic assay on the GeneXpert platform, which is widely available globally but had never been applied to pharmacogenomics. We found that this assay could robustly distinguish wild type, mutant and heterozygous alleles from a range of blood volumes as low as 25 ul, making it suitable for use with venous blood samples or finger-stick blood samples. The assay requires minimal hands-on time for sample preparation, which would facilitate its use in resource-constrained settings.

An earlier model (“NAT2Pred”) predicted NAT2 acetylation phenotype from unphased genomic data; however, it had moderate error rates in distinguishing intermediate from rapid acetylators (33). Moreover, error rates among individuals from Sub-Saharan Africa were 14%, in part due to the exclusion of the G191A (R64Q) SNP, common to the NAT2 *14 allele cluster, which is frequent in Africans and African-Americans, but virtually absent in Caucasian, Indian, and Korean populations (36). A recent study using the set of 6 NAT2 SNPs included in NAT2Pred, but not including G191A, found no correlation between *NAT2* genotype and INH acetylation phenotype in HIV-infected, Zulu individuals with culture-confirmed tuberculosis in Durban, South Africa (37), underscoring the importance of including this SNP in genotype predictions in this region.

Indeed, ethnic differences in SNP frequencies are responsible for the differences in frequency of rapid, intermediate and slow acetylator *NAT2* haplotypes (18, 23). We trained our SNP classifier with globally representative data, which resulted in the selection and inclusion of the G191A SNP in our model and assay. This is particularly important as Sub-Saharan Africa bears a substantial burden of tuberculosis disease and mortality as well as HIV co-infection, which is independently associated with greater pharmacokinetic variability and tuberculosis treatment toxicity (38, 39).

The association between acetylation polymorphisms and INH metabolism was first demonstrated in 1959, and their importance was well characterized in subsequent decades through phenotypic descriptions (40-42). Subsequent genotypic descriptions confirmed that *NAT2* polymorphisms predicted INH early bactericidal activity, and clinical outcomes including hepatotoxicity, relapse and acquisition of drug resistance. Further dosing studies demonstrated that provision of lower doses to slow acetylators and higher doses to rapid acetylators could achieve target concentrations (43). One randomized trial of pharmacogenomic-guided dosing of INH during active tuberculosis treatment found that it significantly reduced toxicities (among slow acetylators) and treatment non-response (among rapid acetylators). Taken together, the evidence for pharmacogenomic guided dosing to achieve consistent drug levels and improve clinical outcomes is strong. Automated, easy-to-use assays could enable pharmacogenomic guided isoniazid dosing in resource constrained settings, where a substantial burden of the world’s tuberculosis occurs.

The findings of this study are subject to several limitations. We tested the assays on 48 individuals with active tuberculosis and 20 healthy individuals with a diverse representation of polymorphisms, but the number of participants with G191A mutations was limited (n=5). A larger validation study involving testing on whole blood, including from finger stick capillary blood, is needed to assess real-world performance of this assay under field conditions. Due to unavailability of whole blood samples from TB patients, we used sputum samples to extract host DNA for genotyping. Further studies should also investigate testing on non-invasive samples including sputum, saliva or oral swabs, from which DNA is abundant. Second, we focused on *NAT2* polymorphisms, as they explain majority of interindividual pharmacokinetic variability, though polymorphisms in several other genes have been associated with hepatotoxicity. However, these associations have been comparatively modest and somewhat inconsistent (44-45). We focused on INH and did not include other important tuberculosis drugs, such as rifampicin. The evidence base for pharmacogenomic markers predicting rifampin pharmacokinetics is less robust, and findings concerning clinical outcomes such as toxicities or treatment response are limited (46-49). However, given the importance of this drug class in treatment of active and latent tuberculosis, and emerging evidence supporting greater efficacy of higher doses of rifampin, further investigation of pharmacogenomic markers in rifampicin is needed. Future assays may include polymorphisms influencing rifampicin metabolism to further optimize treatment of tuberculosis.

Since the demonstration of the efficacy of six month, short-course chemotherapy in 1979, standardized treatment for drug susceptible tuberculosis using weight-based doses has remained essentially unchanged. Additionally, INH remains a major component of regimens for treatment of latent tuberculosis, which is recommended by the WHO for young children, HIV-infected individuals and household contacts of tuberculosis cases (50). More than half the world’s population have slow or rapid acetylation phenotypes, which put them at risk for excessive drug levels resulting in drug toxicities or insufficient drug levels putting them at risk of acquired drug resistance or disease relapse. Dose adjustment based on NAT2 acetylation genotyping can achieve consistent, target drug levels and reduce the incidence of poor clinical outcomes. We developed a prototype automated, cartridge-based assay that can reliably predict acetylation phenotype directly from as low as 25 ul of whole blood. By developing this for the GeneXpert platform, which is widely used in low- and middle-income countries for tuberculosis diagnosis, this assay could make personalized tuberculosis treatment dosing available in resource-constrained settings. Further studies are needed to evaluate its accuracy and clinical impact in real-world clinical settings.

## Data Availability

Data supporting the findings of this manuscript are available in the Supplementary Information files or from the corresponding author upon request.

## Acknowledgements

We thank Veronique Dartois for performing the plasma drug level assays.

## Author contributions

JRA and RV conceived of the study. RV, JC, and JRA designed the experiments. RV, SP, FMM, MTV, ASS and JC collected data. RV, NZ, RS and JRA analyzed data. EW, DG and DP provided technical guidance on the assay development. RV and JRA wrote the first draft of the manuscript, and all authors contributed to the final version.

---

**Supplementary Table 1.** Population information on the dataset used to develop the 5-SNP classifier.

**Supplementary Table 2.** List of primers and probes sequences for multiplex assay with their corresponding fluorophores and quenchers

**Supplementary Table 3.** *NAT2* polymorphism profiles of 48 pulmonary TB patients detected from 5-SNP and 7-SNP genotyping assays. Acetylator phenotypes predicted from 5-SNP, 6-SNP and 7-SNP classifiers and plasma INH and acetyl-INH levels recorded at various time-points.

**Supplementary Table 4. (a)** NAT2 genotype of whole blood samples (n=20) analyzed on 5-plex NAT2-PGx assay and validated on Sanger sequencing. **(b)** NAT2 genotypes of ten whole blood samples analyzed using 200ul, 100ul, 50ul and 25ul samples

## At a Glance Commentary

**Scientific Knowledge on the Subject**

Standardized weight-based dose of isoniazid (INH) may result in variable drug levels in individuals leading to drug-induced toxicities, treatment failure and relapse. Polymorphisms in the *NAT2* gene explain the majority of interindividual pharmacokinetic variability of INH. Pharmacogenomic guided dosing may improve treatment outcomes, but scalable assays are not available.

**What This Study Adds to the Field**

Earlier *NAT2* acetylation prediction algorithms had diminished accuracy in African populations by failing to include a common polymorphism. We developed an algorithm on globally representative data that predicted acetylation genotype with 100% accuracy using 5 unphased SNPs. We tested the accuracy of this acetylator prediction algorithm against INH clearance rates in 48 pulmonary TB patients and found very strong high correlation. We then developed a cartridge-based multiplex assay for these 5 SNPs on the GeneXpert platform that can accurately detect *NAT2* genotype from 25ul whole blood. Because this platform is widely used in low- and middle-income countries for tuberculosis diagnosis, this assay could make personalized isoniazid dosing available in resource-constrained settings.

